# In-Hospital and Long-Term Outcomes in Spontaneous Coronary Artery Dissection (SCAD) with Concurrent Cardiac Arrest: A Systematic Review and Meta-Analysis

**DOI:** 10.1101/2024.09.25.24314406

**Authors:** Omar Baqal, Suganya A. Karikalan, Elfatih A. Hasabo, Haseeb Tareen, Pragyat Futela, Rakhtan K. Qasba, Areez Shafqat, Ruman K. Qasba, Sharonne N. Hayes, Marysia S. Tweet, Hicham Z. El Masry, Kwan S. Lee, Win-Kuang Shen, Dan Sorajja

**Affiliations:** Department of Cardiovascular Medicine, Mayo Clinic, Phoenix, Arizona, USA; CHRISTUS Good Shepherd Medical Center, Longview, Texas, USA; CORRIB Research Centre for Advanced Imaging and Core Laboratory, Clinical Science Institute, University of Galway, Galway, Ireland; Discipline of Cardiology, Saolta Healthcare Group, Health Service Executive, Galway University Hospital, Galway, Ireland; Henry Ford Jackson Hospital, Jackson, Michigan, USA; Department of Cardiovascular Medicine, Mayo Clinic, Rochester, Minnesota, USA; MetroHealth Medical Center, Cleveland, Ohio, USA; Green Life Medical College and Hospital, Dhaka, Bangladesh; College of Medicine, Alfaisal University, Riyadh, Saudi Arabia; Sher-i-Kashmir Institute of Medical Sciences, Srinagar, Jammu & Kashmir, India

**Keywords:** spontaneous coronary artery dissection, cardiac arrest, ventricular arrhythmia, ventricular fibrillation, ventricular tachycardia, sudden cardiac death, myocardial infarction, defibrillation

## Abstract

**Background:** Spontaneous coronary artery dissection (SCAD) is increasingly recognized as an important cause of myocardial infarction (MI). However, our understanding of clinical characteristics that predispose patients to worse outcomes, such as concurrent sudden cardiac arrest (CA), remains limited.

**Objective:** We performed a systematic review and meta-analysis of studies assessing clinical outcomes among SCAD patients with aconcurrent CA.

**Methods:** This study was performed according to PRISMA guidelines. PubMed, Cochrane, and Scopus were systematically searched using relevant search terms, such as “Spontaneous Coronary Artery Dissection”, “Ventricular Tachycardia”, “Ventricular Fibrillation”, “Sudden Cardiac Death” and “Cardiac Arrest”. The search was conducted from database inception to July 2024. Studies assessing the clinical outcomes of SCAD patients with concurrent CA were included. RevMan 5.4 was used for meta-analysis.

**Results:** After removal of duplicates, 269 studies underwent screening, out of which 10 studies were included (n= 3978 patients, 357 with CA). In-hospital mortality, post-discharge mortality, recurrent MI and recurrent SCAD occurred in 20%, 3%, 12% and 9% of SCAD patients with CA, respectively. When compared to SCAD patients without CA, SCAD patients with CA were at significantly higher risk of in-hospital mortality (RR = 6.75, 95% CI [4.50, 10.14], I^2^= 0%), post-discharge mortality (RR = 5.86, 95% CI [1.72, 19.91], I^2^=0%), recurrent MI (RR =3.31, 95% CI [2.03, 5.39], I^2^=56%), recurrent SCAD (RR = 1.91, 95% CI [1.11, 3.27], I^2^= 43%), acute heart failure (RR = 4.82, 95% CI [3.22, 7.20], I^2^=42%), and cardiogenic shock (RR = 6.11, 95% CI [4.07, 9.19], I^2^=64%). Out of a pooled 24 implanted cardiac defibrillators (ICDs) and 11 wearable cardiac defibrillators (WCDs), there was only one appropriate and one inappropriate ICD discharge recorded over the follow-up period.

**Conclusion:** In this cohort, SCAD with concurrent CA was associated with worse in-hospital and long-term outcomes including in-hospital and post-discharge mortality, acute heart failure, and recurrent MI and SCAD. There was a low long-term rate of administered defibrillator therapies among patients discharged from the hospital with an ICD or WCD. Further research is needed to better delineate optimal management approaches toward this high-risk patient population, including secondary prevention of sudden cardiac death.

**Graphical abstract:** 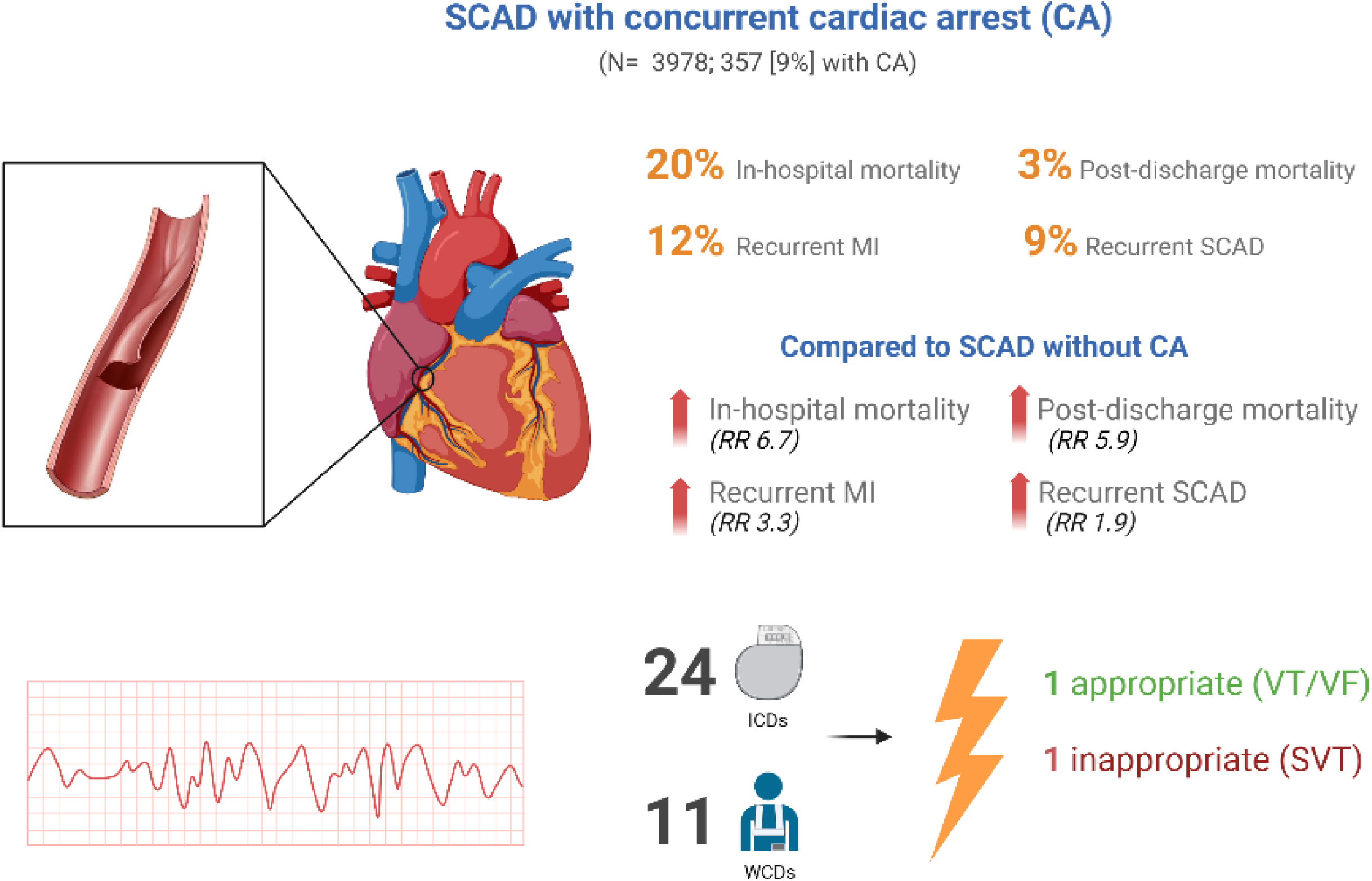

## Introduction

Spontaneous coronary artery dissection (SCAD) is a form of acute coronary syndrome (ACS) characterized by the non-traumatic, non-iatrogenic intimal tear or separation of the coronary arterial tunica intima and media by an intramural hematoma ^1,2^. This is often in the setting of a pre-existing arteriopathy weakening the arterial wall, with additional precipitating emotional or physical stressors that incite the dissection ^3–6^. Although its true prevalence is unknown, the increasing use of advanced intracoronary imaging techniques and greater awareness has led to SCAD being recognized as an important etiology of ACS. A disease of young-to-middle aged women, SCAD is responsible for 24-35% of cases of ACS in women < 60 years of age ^7^. Predisposing conditions to SCAD include fibromuscular dysplasia, inheritable connective tissue diseases, and pregnant/postpartum status^3,4^. The most common presentation of SCAD is non-ST segment elevation myocardial infarction (NSTEMI) followed by STEMI, with an estimated 10-20% presenting with cardiac arrest ^8^.

The presence of ventricular arrhythmia (VA) has been reported in 3-13% of patients with SCAD and has been associated with worse in-hospital and long-term outcomes, including mortality and MACE^9^. Risk stratification for sudden cardiac death (SCD) in SCAD remains poorly explored, and guidelines are unclear on whether or not to pursue ICD implantation in patients with a reversible cause of SCD ^9^. Additionally, data remains scarce pertaining to optimal revascularization strategies, timing of ICD implantation, long-term ICD outcomes, and role of wearable cardiac defibrillators (WCDs) at hospital discharge in patients with SCAD and concurrent CA.

To consolidate published literature on the topic, we performed a systematic review to explore the impact of concurrent CA in SCAD on in-hospital and long-term clinical outcomes.

## Methods

This meta-analysis was carried out and reported per the Preferred Reporting Items for Systematic Reviews and Meta-analyses (PRISMA) ^10^. The study was registered on PROSPERO with registration ID CRD42024511286.

### Literature search

We performed a search for studies reporting outcomes of people with SCAD complicated by CA, which included VT/VF, pulseless electrical activity (PEA) and asystole. Searches were performed on PubMed/MEDLINE,

Cochrane, and Scopus using keywords and standardized index terms, such as “Spontaneous Coronary Artery Dissection”, “Ventricular Tachycardia”, “Ventricular Fibrillation”, “Sudden Cardiac Death” and “Cardiac Arrest”. The search was conducted from date of database establishment to July 2024.

### Inclusion and exclusion criteria for identification of studies

Articles were considered eligible for inclusion if they were written in English, were peer-reviewed observational studies or randomized clinical trials; included adults ≥ 18 years of age, reported on outcomes of patients with SCAD and concurrent CA, and included information on hospital follow-up and longer. Studies that did not report on the distinct outcomes of SCAD with CA, included participants under 18 years of age, or those with no hospital or post-discharge follow-up were excluded. We also excluded meta-analyses, systematic reviews, case reports, abstracts, editorials, commentaries, and letters to the editor.

### Data screening

Two reviewers (SK and HT) screened the title and abstracts of all retrieved studies using the pre-defined selection criteria. The full texts of studies meeting the criteria were reviewed by OB and RQ. Any disagreements regarding article inclusion were resolved after discussing with the senior reviewer (DS).

### Data extraction

Two authors (HT and OB) extracted data simultaneously into a standard data extraction sheet. The data collected included in-hospital overall mortality, post-discharge overall mortality, major adverse cardiovascular events (MACE) acute heart failure, cardiogenic shock, LVEF (%), recurrent MI, and recurrent SCAD which predominantly included recurrent de novo SCAD. Post-discharge mortality was defined as mortality after hospital discharge over follow-up period. Cardiac arrest either on presentation (including out-of-hospital cardiac arrest) or index in-hospital cardiac arrest was considered. In-hospital and long-term recurrent MI and SCAD on follow-up were included. Any discrepancies were resolved after consulting the senior reviewer (DS).

### Assessment of methodological quality and risk of bias

The reviewers then assessed the risk of bias for each study using the Newcastle-Ottawa Scale (NOS) for prospective and retrospective cohort studies, and adapted NOS for studies without control arm (Total score is 6). The NOS rates observational cohort studies based on the selection of participants, comparability between the exposed and unexposed groups, and the assessment of the association between exposure and outcome. Studies with a score <5 are considered to be of low quality. All included studies scored ≥ 5. **(Supplementary material)**

### Statistical analysis

Data were extracted, and the results were presented as risk ratio (RR) and pooled percentage. For single-arm meta-analysis; we applied the formula: SE = √p (1-p)/ n, where p stands for prevalence to calculate SE. Heterogeneity was assessed using the *I*^2^ test, which calculates percentage variability in endpoints that is due to heterogeneity rather than to chance. The larger the *I*^2^, the more likely that any observed statistical outcomes are due to heterogeneity. An *I*^2^ *<*30% for an outcome of interest was set as the standard for low heterogeneity. All statistical tests were 2-sided, and a *P-*value <0.05 was considered statistically significant. We used the random-effects model in the presence of significant heterogeneity (p <0.1). The statistical analysis was performed using RevMan 5.4. The primary outcome of our study was to compare in-hospital overall mortality between SCAD patients with and without CA. Secondary outcomes of interest included post-discharge overall mortality, acute heart failure, cardiogenic shock, recurrent MI, recurrent SCAD, and LVEF ≤ 40%. Finally, to assess for publication bias, we constructed funnel plots to assess asymmetry for each effect estimate and Egger’s test as a statistical test of funnel plot asymmetry. **(Supplementary material)**

## Results

### Search results and study characteristics

A total of 269 non-duplicate study abstracts and titles were screened. Following the pre-defined exclusion and inclusion criteria, 10 studies were selected for inclusion (**Figure 1**) ^5,9,11–17^. Detailed study characteristics are outlined in **Table 1**. Out of 10 included studies, two were prospective ^13,15^, and eight were retrospective ^5,9,12,14,16–19^. The studies represented various geographical regions, with four studies from the United States ^5,9,12,14,19^, two from Canada ^13,15^, one from Switzerland ^11^, one from Italy ^17^, one from Italy and Spain ^18^, and one from the Middle East (Kingdom of Saudi Arabia, United Arab Emirates, Kuwait, and Bahrain) ^16^. A total of 3978 SCAD patients were included, out of which 357 (9%) had concurrent CA. In addition to VT/VF, Giacobbe et al. ^18^ and Phan et al. ^14^ included PEA and asystole in their definition of cardiac arrest. Two studies reported CA on presentation, three studies reported CA on presentation and during hospitalization, and five studies did not specify CA timing. Six studies included SCAD confirmed by coronary imaging, including angiography. Daoulah et al. (51%), Krittanawong et al (64%) and Tan et al. (78%) reported the lowest proportions of SCAD patients of female sex. Pooled patient sample was predominantly of female sex and <65 years of age.

**Figure 1:**
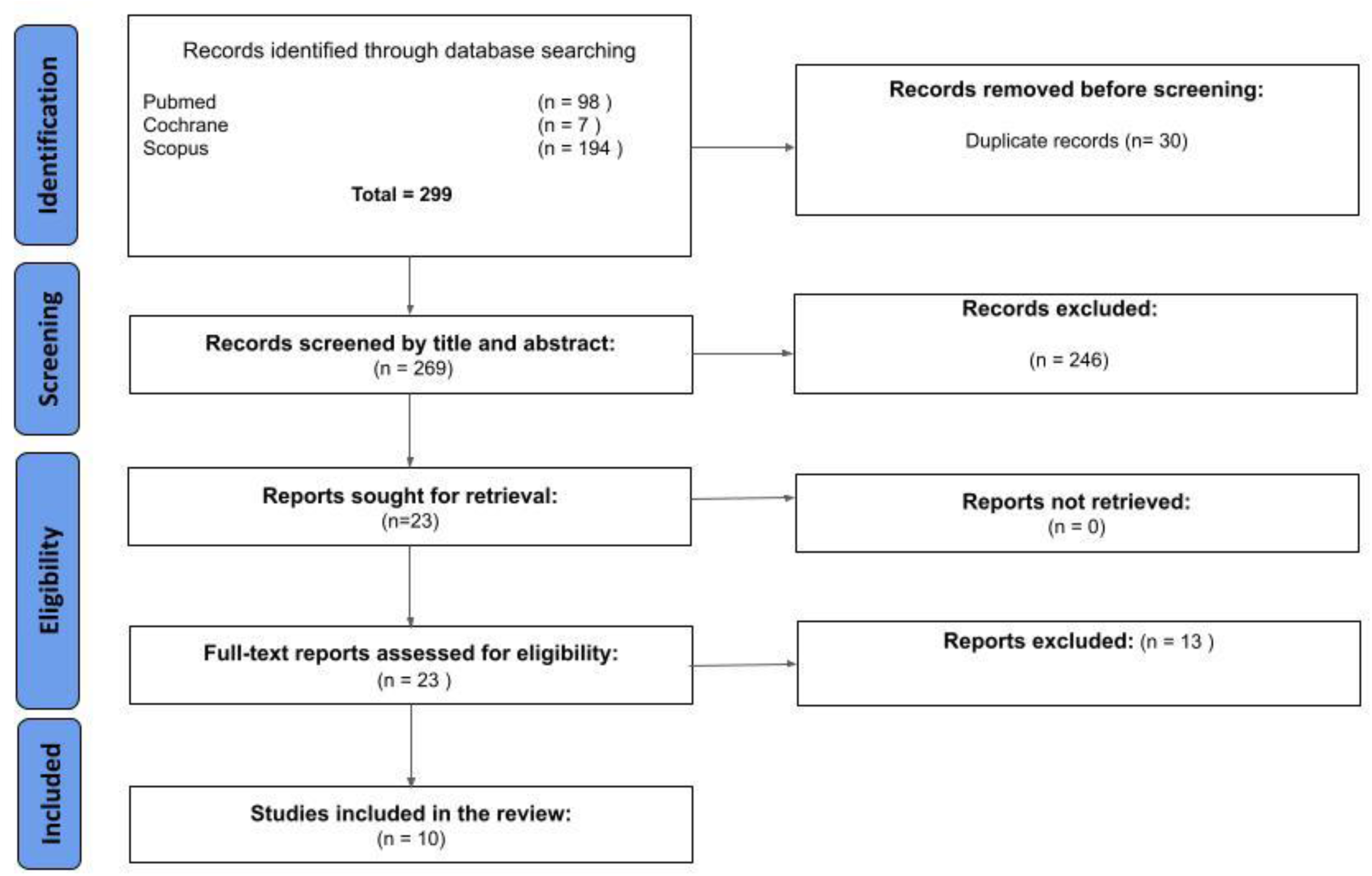
Flow diagram of literature search and selection of publications.

**Table 1:** Characteristics of included studies.

| Study | Type of Study | Definition of CA | Total no. of SCAD patients | Patients with CA (n, %) | CA on presentation (n, % of total CA) | Follow-up duration (months) | Duration of hospital stay (days) | Angiographic confirmation of SCAD? (Y/N) | Age (years) | Female n (%) | Defibrillator therapies | Defibrillator outcomes | SCAD type (% overall) ¶ | Overall revascularization (n, %) |
| --- | --- | --- | --- | --- | --- | --- | --- | --- | --- | --- | --- | --- | --- | --- |
| **Giacobbe 2024**<sup>18</sup> | Retrospective | PEA, asystole, VF, pulseless VT | 375 | 20 (5) | 6 (30) at presentation<br>14 (70) in-hospital | 21 (10.7–46) (median) | 7.5 ± 0.49 | Y | 52.6 ± 10.2 | 315 (84) | 1 ICD<br>2 WCDs | - | Type 1: 23%<br>Type 2: 50%<br>Type 3: 6%<br>Type 4: 26% | PCI: 130 (35%)<br>CABG: 1 (0.2%) |
| **Tan 2023**<sup>12</sup> | Retrospective (NRD) | ICD-10 codes for VT and VF | 877 | 118 (13) | Not specified | 1 | 7.8 ± 8.3 | N | 52.6 ± 13.8 | 687 (78) | - | - | - | - |
| **Antonutti 2021**<sup>17</sup> | Retrospective | Sustained VT/VF | 70 | 7 (10) | Not specified | 39.1 (IQR 13.9 - 86.6) (median) | - | Y | 52 (47-58) | 60 (86) | - | - | Type 1: 56%<br>Type 2: 43%<br>Type 3: 1% | PCI: 13%<br>CABG: 3% |
| **Cheung 2020**<sup>13</sup> | Prospective | Sustained VT/VF | 1056 | 84 (8) | Not specified | 57.6 ± 39.6 (mean) | - | Y | 49.3 ± 11.3 | 941 (89) | 8 ICDs | 1 shock for VT/VF | - | Overall: 174 (16.5%) |
| **Phan 2020**<sup>14</sup> | Retrospective | Cardiac arrest, including PEA, asystole, VT/VF | 208 | 11 (5) | Not specified | 56.4 ± 37.2 (mean) | - | Y | 48.53 ± 12.06 | 186 (94) | 2 ICDs<br>1 WCD | 0 shocks | - | PCI: 23 (11%)<br>CABG: 9 (4.6%) |
| **Krittanawong 2020**<sup>5</sup> | Retrospective | Not specified | 375 | 20 (5) | Not specified | - | - | N | 52.2 ± 12.8 (overall) | 241 (64) | - | - | - | - |
| **Daoulah 2020**<sup>16</sup> | Retrospective | Sustained VT/VF | 83 | 10 (12) | 10 (100) | 18.8 (IQR 9.0-40.1) (median) | - | Y | 46 (26–81) | 42 (51) | 1 ICD | 1 death confirmed 10 months post-discharge, unknown cause | Type 1: 52%<br>Type 2: 42%<br>Type 3: 4% | Overall: 60%<br>PCI: 53%<br>CABG: 7% |
| **Chen 2020**<sup>9</sup> | Retrospective | VT/VF | 349 | 20 (6) | 17 (85) | 69.6 ± 54 (mean) | - | Not specified | 47 ± 12 | 19 (95) for CA group | 5 ICDs 4 WCDs | 0 shocks | - | PCI: 7 (2%) |
| **Saw 2017**<sup>15</sup> | Prospective | VT/VF | 327 | 29 (9) | 29 (100) | 37.2 (IQR 17.9 - 65.9) (median) | 3 (IQR 2-5) (median) | Y | 52.5 ± 9.6 (overall) | 297 (91) | 9 “cardioversion or ICD” | - | Type 1: 26%<br>Type 2: 70%<br>Type 3: 5% | Overall: 61 (18.7%)<br>PCI: 16.5%<br>CABG: 2.1% |
| **Sharma 2017**<sup>19</sup> | Retrospective | Not specified | 102 | 14 (14) | 7 (50) OOH<br>4 (29) in ED<br>3 (21) during/shortly after PCI | 42 (mean) | - | Not specified | 44 ± 11 | 88 (86) | 7 ICDs<br>4 WCDs | 1 shock for SVT | Not specified | PCI: 37 (36%)<br>CABG: 3 (3%) |

CA: Cardiac arrest, NRD: National Readmissions Database, OOH: Out of hospital, ED: Emergency department, PCI: percutaneous coronary intervention, SVT: Supraventricular tachycardia
\* Data cited is for SCAD patients with CA unless specified otherwise
¶ Saw J. Coronary angiogram classification of spontaneous coronary artery dissection. *Catheter Cardiovasc Interv.* 2014;84(7):1115-1122.

### Clinical outcomes

A single-arm meta-analysis was performed to assess the prevalence of study endpoints among SCAD patients with concurrent CA. A pooled analysis revealed that in-hospital mortality, post-discharge mortality, recurrent MI and recurrent SCAD occurred in 20%, 3%, 12% and 9% of SCAD patients with CA, respectively. **(Figure 2).** Out of a pooled 24 ICDs and 11 WCDs, there was only one appropriate ICD discharge reported for VT/VF, and one inappropriate ICD discharge for supraventricular tachycardia (SVT) **(Table 1).**

**Figure 2:**
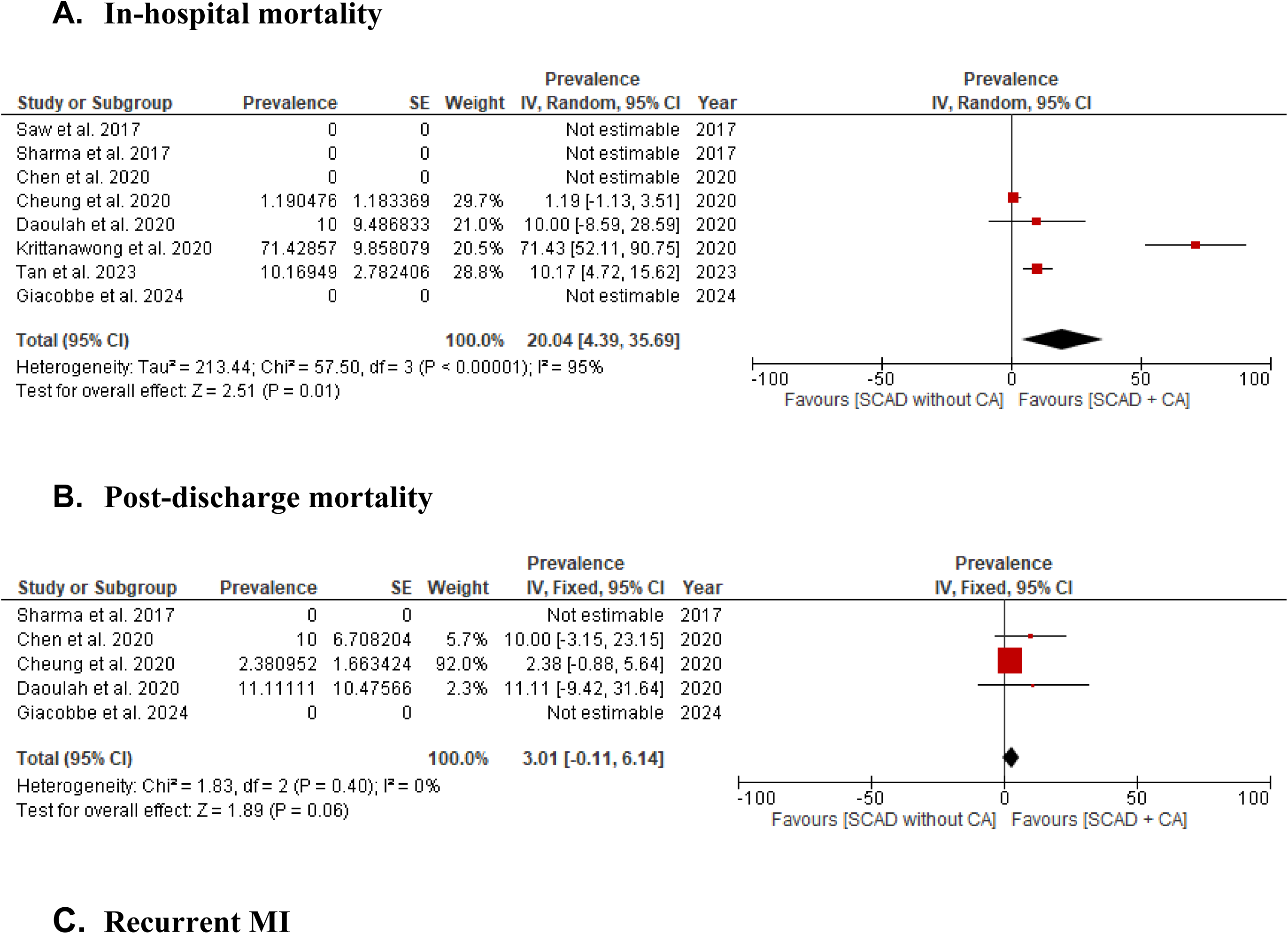
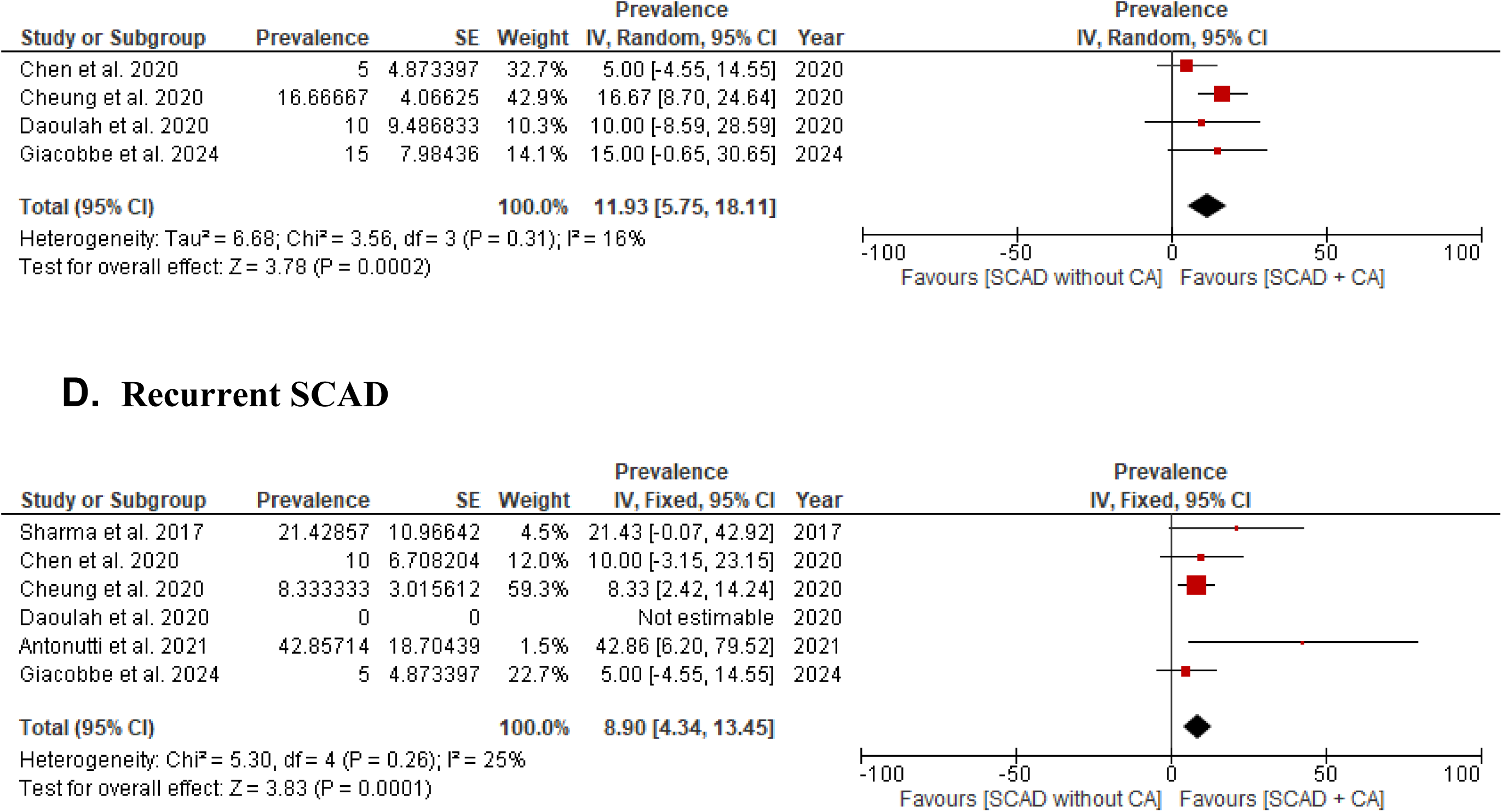
Outcomes among SCAD patients with CA compared to SCAD patients without CA.

### In-hospital overall mortality

Seven studies were included in the meta-analysis and the pooled results showed that SCAD patients with CA had higher in-hospital overall mortality compared to SCAD patients without CA (27/295 vs 47/2900, RR = 6.75, 95% CI [4.50, 10.14], P < 0.00001, I^2^= 0%). (Figure 3A)

**Figure 3:**
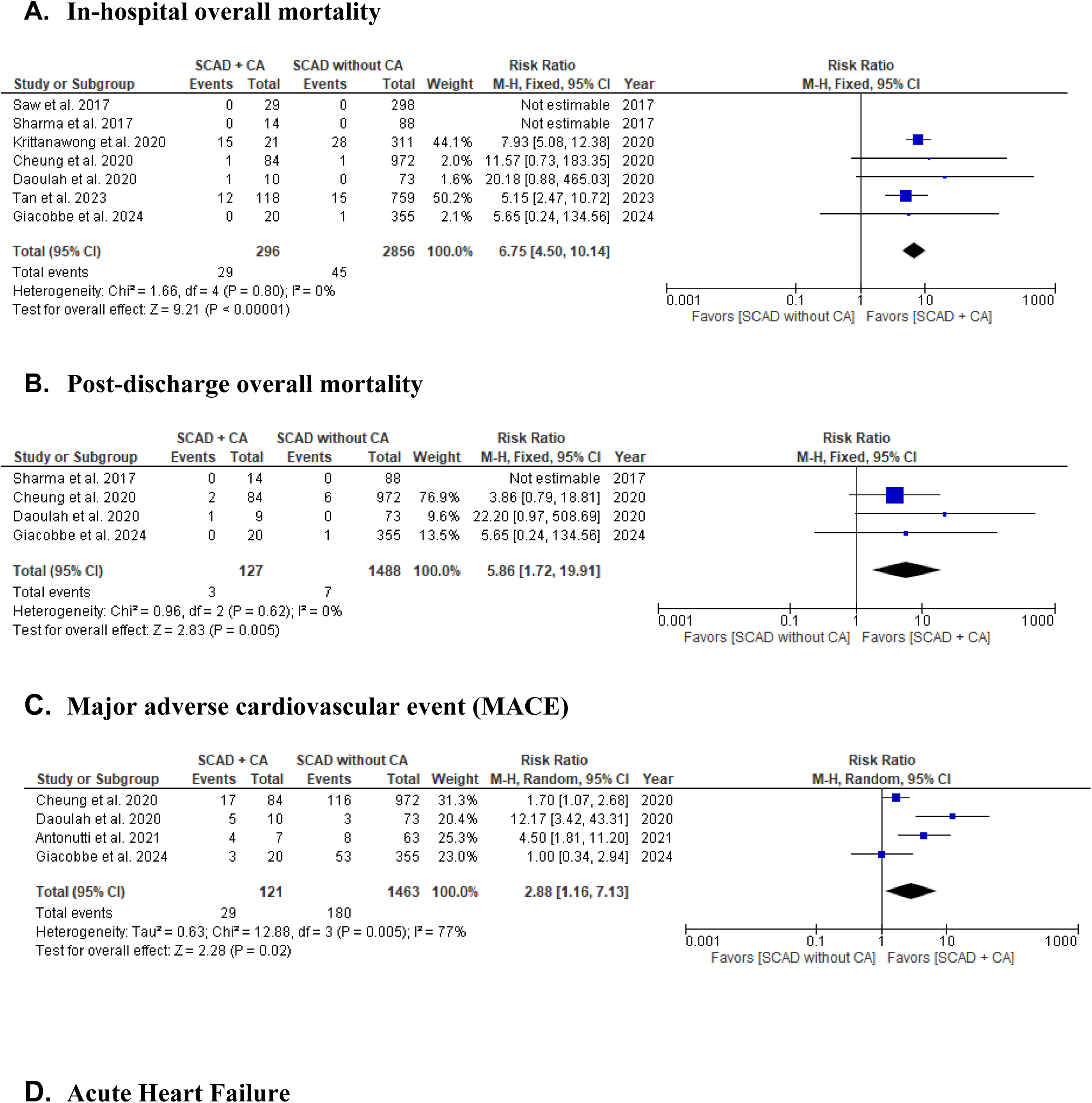
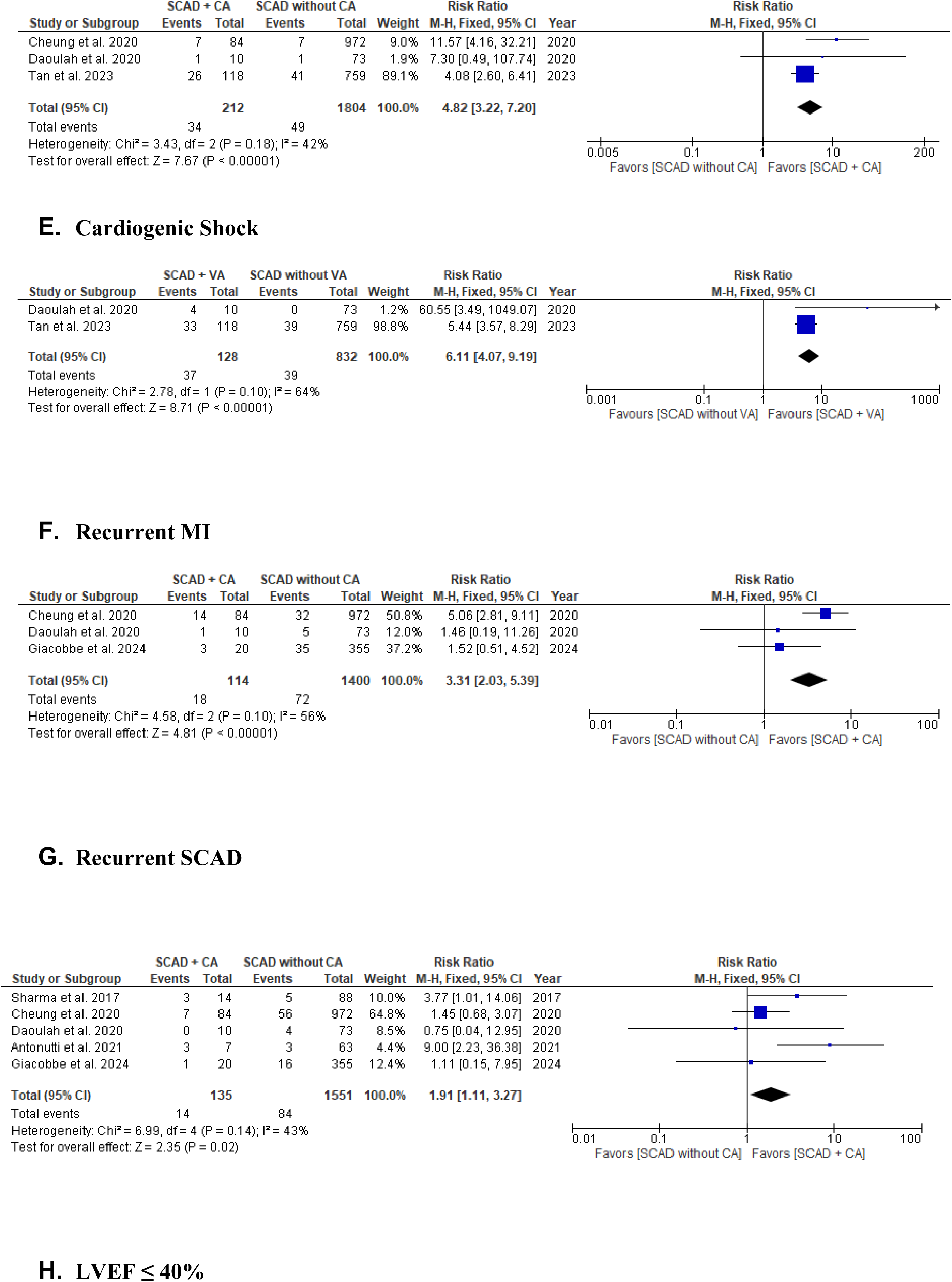
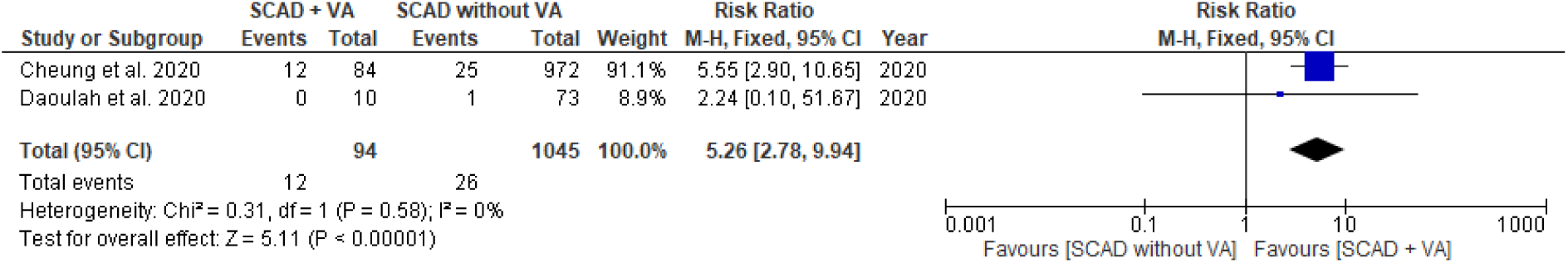

### Post-discharge overall mortality

Four studies were included in the meta-analysis. Follow-up ranged from 18.8 to 39.1 months for studies reporting median follow-up and 42.0 to 69.6 months for studies reporting mean follow-up. NRD-based study by Tan et al. assessed 1-month post-discharge outcomes. The pooled results showed that SCAD patients with CA had higher post-discharge overall mortality compared to SCAD patients without CA (3/127 vs 7/1488, RR = 5.86, 95% CI [1.72, 19.91], P = 0.005, I^2^=0%). (Figure 3B)

### Major adverse cardiovascular events (MACE)

Four studies were included in the meta-analysis and the pooled results showed that SCAD patients with CA had higher risk of MACE compared to SCAD patients without CA (29/121 vs 180/1463, RR = 1.91, 95% CI [1.11, 3.27], P=0.02, I^2^=77%). High heterogeneity was noted, potentially attributable to varying definitions of MACE across studies. (Figure 3C)

### Acute heart failure

Three studies were included in the meta-analysis and the pooled results showed that SCAD patients with CA had higher risk of acute heart failure compared to SCAD patients without CA (34/212 vs 49/1804, RR = 4.82, 95% CI [3.22, 7.20], P < 0.00001) (I^2^= 42%). (Figure 3D)

### Cardiogenic shock

Two studies were included, and the pooled results showed that SCAD patients with CA had higher risk of cardiogenic shock compared to SCAD patients without CA (37/128 vs 39/832, RR = 6.11, 95% CI [4.07, 9.19], P < 0.00001) (I^2^= 64%). (Figure 3E)

### Recurrent MI

Three studies were included, and the pooled results showed that SCAD patients with CA had higher risk of recurrent MI compared to SCAD patients without CA (16/114 vs 134/1400, RR =3.31, 95% CI [2.03, 5.39], P < 0.00001), I^2^=56%) (Figure 3F)

### Recurrent SCAD

Recurrent SCAD primarily included recurrent de novo SCAD, and not SCAD extension. Five studies were included, and the pooled results showed no significant difference in SCAD recurrence between SCAD patients with CA compared to SCAD patients without CA (14/135 vs 84/1551, RR = 2.48, 95% CI [0.59, 10.47], P = 0.22) (I^2^= 65%). (Figure 3G)

### LVEF ≤ 40%

Three studies were included, and the pooled results showed that SCAD patients with CA had higher risk of LVEF ≤ 40% compared to SCAD patients without CA (20/105 vs 52/1242, RR = 5.26, 95% CI [2.78, 9.94], P < 0.00001) (I^2^= 0%). (Figure 3H)

## Discussion

We present, to the best of our knowledge, the largest analysis on the clinical outcomes in SCAD with concurrent cardiac arrest. Patients with SCAD and CA were at a higher risk of in-hospital and post-discharge mortality, acute heart failure, cardiogenic shock, recurrent MI and recurrent SCAD. Importantly, our results are mostly applicable to SCAD patients with CA who survive at least up to hospital arrival to undergo medical evaluation and additional testing. Those who do not survive to hospital presentation may have unique clinical characteristics and risk factors that are likely largely understudied.

Although SCAD is generally associated with good long-term outcomes with low overall mortality and incidence of MACE ^4^, our pooled double-arm meta-analysis highlights that patients who develop concurrent CA be considered high-risk for both acute and long-term complications. Nevertheless, cause-specific mortality in these patients is not well explored. Patients with SCAD and CA are more likely to present with STEMI, which is commonly associated with larger infarct size and increased risk of LV dysfunction and adverse cardiovascular outcomes ^9^. Krittanawong et al. reported that atrial fibrillation, steroid use, ventricular arrhythmias, and cardiac arrest were independent predictors of in-hospital mortality in SCAD patients ^5^. Conservative management at index presentation is typically pursued, although Waterbury et al. highlighted that 17.5% of conservatively managed SCAD patients had early SCAD progression based on angiographic evaluation; specifically, patients with intramural hematoma at baseline were at higher risk of progression and early deterioration ^20,21^.

Studies on the community-based Canadian SCAD cohort ^4^, the Nationwide Readmissions Database ^12^, the Mayo Clinic SCAD Virtual Multi-Center registry ^22^, and a multinational study covering four Gulf countries (KSA, UAE, Kuwait, and Bahrain) ^16^ have recorded rates of CA between ∼3-13% at index hospitalization and ∼4% of patients during post-discharge follow-up, corresponding with our pooled estimates. The reported prevalence of CA in SCAD patients is more variable, which could be explained by a multitude of reasons, including baseline patient features and SCAD severity, and time frame over which CA was assessed (e.g., immediate post-event, in-hospital, and long-term follow-up).

Management recommendations for SCAD are based on observational studies, and there are no published randomized control trials yet. Consensus statements by the European Society of Cardiology (ESC) and American Heart Association (AHA) on SCAD recommend medical management when possible for SCAD patients who are hemodynamically stable and do not show signs of ongoing myocardial ischemia ^2,23^. Long-term management of SCAD involves maintenance on aspirin and beta-blockers, the latter associated with reduced incidence of SCAD recurrence in one observational study ^15^. This guidance is based on findings that the dissection will most likely heal spontaneously, and recurrence typically does not involve the initial culprit coronary artery, nor does PCI prevent future SCAD recurrence ^21,24^. In our meta-analysis, only three studies had data on beta-blocker usage, although specific data on SCAD patients with concurrent CA was lacking, and use of antiarrhythmic medications was unavailable^11,15,17^. In a prospective, multicenter study by Saw et al. of 750 SCAD patients out of whom 8.3% had VT/VF, aspirin and beta-blocker usage was high with low long-term mortality ^4^. Tweet et al. reported an uncomplicated hospital course among 31 SCAD patients managed conservatively, with two deaths occurring on long-term follow-up. In contrast, failure rate of PCI was 53% and it did not significantly reduce rates of revascularization and recurrence of SCAD ^24^. One population very much underrepresented in the included studies is obstetric patients; pregnancy and the peri-partum period have been posited as major risk factors for SCAD in multiple studies with a more severe presentation ^25^. Among 13 deaths attributable to pregnancy-associated SCAD (P-SCAD) in the MBRRACE-UK audit, 12 suffered from an out-of-hospital CA ^26^. Phan et al. found that SCAD patients with CA were more likely to have P-SCAD ^14^. However, Krittanawong et al. in their analysis based on NRD data noted a similar incidence of CA between P-SCAD and non-P-SCAD ^27^. Therefore, further research is needed to clarify possible risk-enhancing factors for CA and CA-related outcomes in this patient group.

Those who receive PCI show no statistical difference in long-term outcomes compared to those who received no PCI, and the former may expose patients to greater risks of iatrogenic complications such as propagation of intramural hematoma and dissection and abrupt vessel occlusion ^4,24,28,29^. This can lead to an increased myocardial ischemic burden and subsequently increased risk of ventricular arrhythmogenicity. Nearly half of the VT/VF arrests reported in their cohort by Phan et al. occurred during PCI ^14^. A meta-analysis by Bocchino *et al.* also showed that the mean success rate of PCI was less than 50% and that revascularization therapies did not decrease rates of all-cause mortality, cardiovascular mortality, myocardial infarction, heart failure, and SCAD recurrence ^30^. In light of uncertain benefits and increased risk of bleeding complications associated with antiplatelet therapy in SCAD patients who do not undergo PCI, experts recommend individualized decision-making considering the risk-benefit ratio ^2^. Indications for the use of more advanced imaging modalities like intravascular ultrasound (IVUS) and optical coherence tomography (OCT) are also not well defined but may optimize the use of PCI in high-risk patients, though their uptake in studies has been low ^30,31^.

Notably, based on our pooled sample which included placement of 24 ICDs and 11 WCDs, there was only one appropriate ICD discharge reported for VT/VF by Cheung et al. over a mean follow-up of 4.8 ± 3.3 years, and one inappropriate discharge for SVT ^13,19^. In a community-based study of 27 patients with VA due to SCAD, Chen *et al.* demonstrated that no patient developed recurrent VA due to SCAD and none required defibrillation therapies over a 5.8-year mean follow-up ^9^. Over a mean follow up period of 3.5 years following ICD placement among seven SCAD patients with SCD as part of the MGH-SCAD registry, there was one inappropriate ICD discharge for supraventricular tachycardia ^19^. The unclear risk-benefit ratio of ICD placement for secondary SCD prevention in SCAD was highlighted by Garg et al in their meta-analysis, noting 1.2% (95% CI: 0%–15.8%, I2 = 0%) and 1% (95% CI: 0%–15.3%, I2 = 0%) patients received appropriate and inappropriate ICD therapies, respectively, during the follow-up period ^32^. Given these findings and considering that most SCAD lesions heal spontaneously on follow-up, current guidelines do not support early ICD implantation for patients after an episode of cardiac arrest with a reversible cause ^33,34^. Our results support the idea of SCAD-mediated CA being a transient substrate with a low recurrent event rate, as long as the LVEF recovers. Factors such as incomplete coronary revascularization, persistently reduced LVEF, and recurrent VA can also be used to risk-stratify sudden cardiac death and guide decision-making regarding ICD placement ^35^. It would be of value for future studies to stratify patient outcomes data based on timing of CA – such as out-of-hospital, in-hospital and procedural. Data remains scarce on use of multi-modality imaging for determining ICF placement, optimal timing of ICD placement, long-term appropriateness of ICD therapies in this patient population and role of WCDs as a bridge to ICD implantation, and further research is needed.

We also found that the incidence of recurrent de novo SCAD was higher in SCAD patients with CA. Management of modifiable factors such as smoking cessation, avoidance of pregnancy, and adherence to long-term medical therapy with beta-blockers may decrease the risk of recurrent SCAD. The development of VA in SCAD patients can be related to multiple factors, with only one of them being recurrent SCAD. For instance, myocardial fibrosis after SCAD-induced MI may also increase the long-term risk of VAs. Hence, future studies evaluating the association between myocardial fibrosis as assessed by imaging techniques such as cardiac MRI (CMR) and recurrent VAs may reveal new strategies for identifying candidates that may benefit from ICD placement.

Indeed, it is well recognized that solely relying on LVEF is inadequate in stratifying SCAD patients’ future risk of SCD; the cut-off of LVEF < 35% that guides ICD placement may correctly predict only ∼20% of SCD events in other cardiomyopathies ^36^. CMR—particularly late gadolinium enhancement (LGE) to detect myocardial fibrosis— provides additional prognostic information about SCD risk over solely LVEF ^37–39^. Replacement of viable myocardial tissue by fibrosis in SCAD is likely to significantly enhance risk of post-SCAD arrhythmias. Hence, characterizing such an arrhythmogenic substrate by CMR-LGE may enhance SCD risk stratification and further guide appropriateness of ICD implantation. In this context, one study of 14 patients with acute or recurrent SCAD demonstrated LGE in the distribution of the culprit coronary artery in all cases ^40^. This finding needs to be replicated in larger cohorts. Patient populations who would benefit most from CMR-based characterization of scar tissue require further defining. For instance, Androulakis *et al.* demonstrated that SCAD involving the RCA was predictive of myocardial fibrosis detected by LGE (OR = 5.2; p = 0.034)^41^. Additionally, most patients with SCAD may not present with large infarct sizes and subsequent myocardial fibrosis; however, those with pregnancy-related SCAD, multi-vessel SCAD, STEMI, CA, extracoronary arteriopathies (*e.g.,* fibromuscular dysplasia, focal stenoses, and arterial tortuosity) are more likely to present with larger infarct sizes and may therefore benefit from further anatomical and functional characterization by CMR and LGE ^41,42^.

### Limitations

All included studies were observational in nature, with results impacted by the completeness of collected and analyzed data. Tan et al. was a database study utilizing the National Readmissions Database and used ICD coding for coronary artery dissection due to lack of specific ICD-10 codes for SCAD, possibly contributing to heterogeneity of studied patients due to potential inclusion of iatrogenic coronary artery dissection and atherosclerotic coronary artery dissection. Similarly, Krittanawong et al. was an ICD-9 or -10 codes-based study without angiogram confirmation which may contribute to heterogeneity. These studies had a higher proportion of men, suggesting that some of the patients may have had dissection in the setting of atherosclerosis. As our study was a systematic review and meta-analyses of published observational studies, some results may be hindered by publication biases.

Heterogeneity was noted among several variables, which can be attributed to differences in study populations, definitions of endpoints, and other clinical and/or procedural variables. Differences in population characteristics, interventions, and outcomes measured can create discrepancies that make it challenging to draw generalized conclusions. Similarly, these cohorts only included patients who survived long enough to be diagnosed with SCAD and did not include autopsy data. SCAD is a particularly important clinical entity among obstetric patients, although our review noted data on this patient group to be lacking and so was literature on sex-based differences in outcomes. Further research is needed to explore outcomes in women and obstetric patients with SCAD and CA. Due to limitations in the reported literature, our study did not focus on sex-based differences or pregnancy associated SCAD. Although our study is, to the best of our knowledge, the largest to study clinical outcomes in SCAD patients with CA, it is important to acknowledge that these findings are hypothesis-generating and may not be powered sufficiently to guide clinical care in a diverse patient population with varying characteristics and risk factors.

### Conclusions

Our systematic review and meta-analyses highlight that concurrent CA in SCAD is associated with worse in-hospital and post-discharge mortality, and higher risk of acute heart failure, cardiogenic shock, recurrent MI and recurrent SCAD. Pooled follow-up ICD and WCD data noted a low recurrent CA event rate. Our findings support SCAD- mediated CA to be a transient substrate, supporting a conservative approach to secondary CA prevention in SCAD consistent with the current recommendation for atherosclerotic CAD/MI-mediated CA. Further research is needed on various aspects of management relevant to this high-risk patient population, including patient selection for ICD placement and the role of WCDs in managing early recurrent SCD.

### Disclosures

Dr. Marysia Tweet is supported by a grant from the National Heart, Lung, and Blood Institute, NIH K23H155506. The Mayo Clinic “Virtual” Multicenter SCAD Registry receives support from SCAD Research, Inc.

## Data Availability

No new original data generated. Data utilized in current study available through manuscripts and supplemental material of included studies.

## Acknowledgements

Graphical abstract created with BioRender. Blood vessel image in graphical abstract used with permission of Mayo Foundation for Medical Education and Research, all rights reserved.

## Supplementary material

**Supplementary table 1:** Quality assessment of included studies using adapted Newcastle-Ottawa Scale (NOS)

| Study | Selection | Comparability | Outcome | Total score |
| --- | --- | --- | --- | --- |
| Giacobbe et al. 2024 | 3 | 1 | 3 | 6 |
| Antonutti et al. 2021 | 3 | - | 3 | 6 |
| Tan et al. 2023 | 3 | 1 | 3 | 7 |
| Cheung et al. 2020 | 4 | 0 | 3 | 7 |
| Krittanawong et al. 2020 | 4 | 0 | 1 | 5 |
| Phan et al. 2020 | 4 | 1 | 2 | 7 |
| Saw et al. 2017 | 3 | - | 2 | 5 |
| Chen et al. 2020 | 3 | - | 2 | 5 |
| Daoulah et al. 2020 | 3 | 1 | 2 | 6 |
| Sharma et al. 2017 | 3 | 1 | 2 | 6 |

**Supplementary Figure 2:**
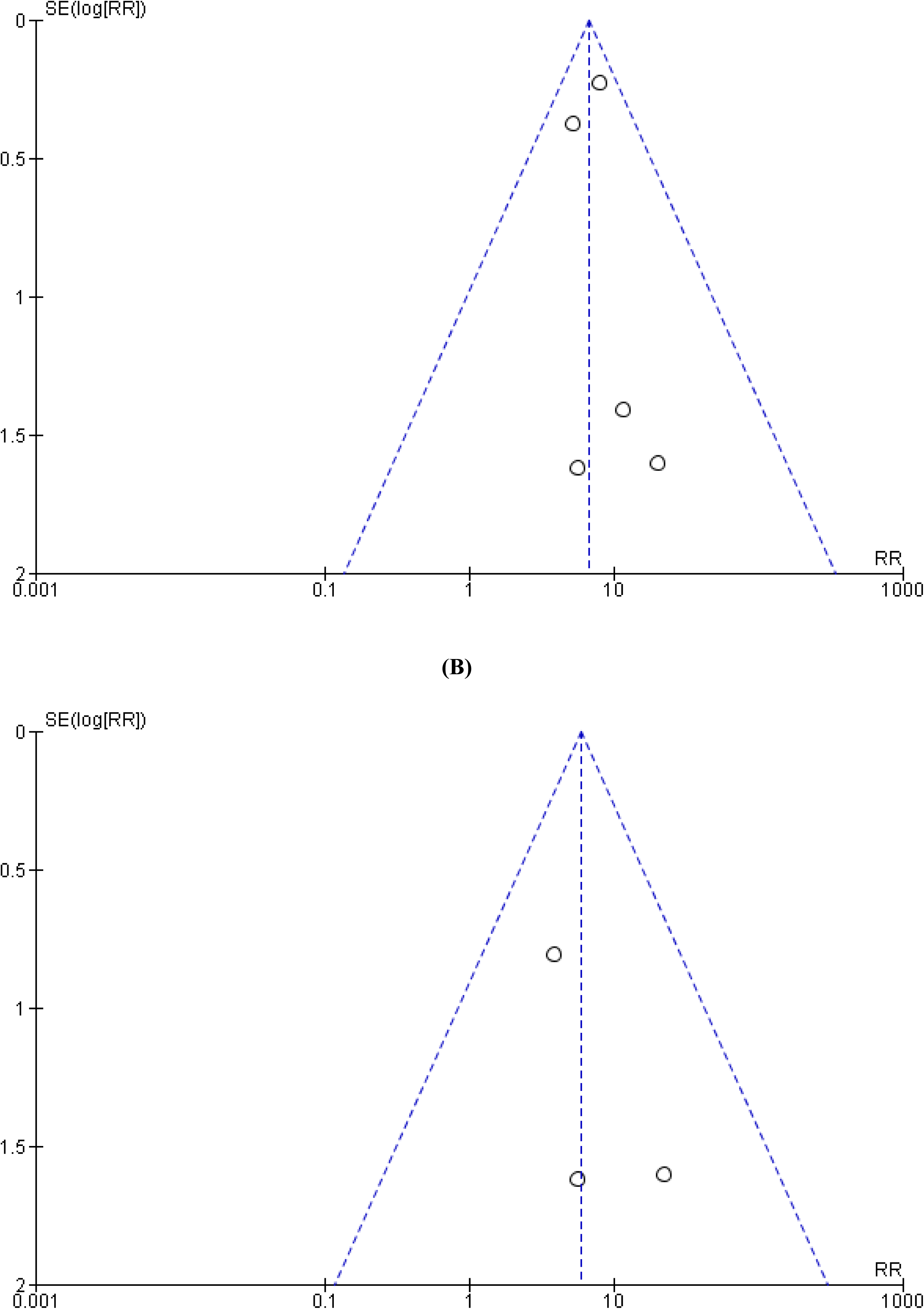
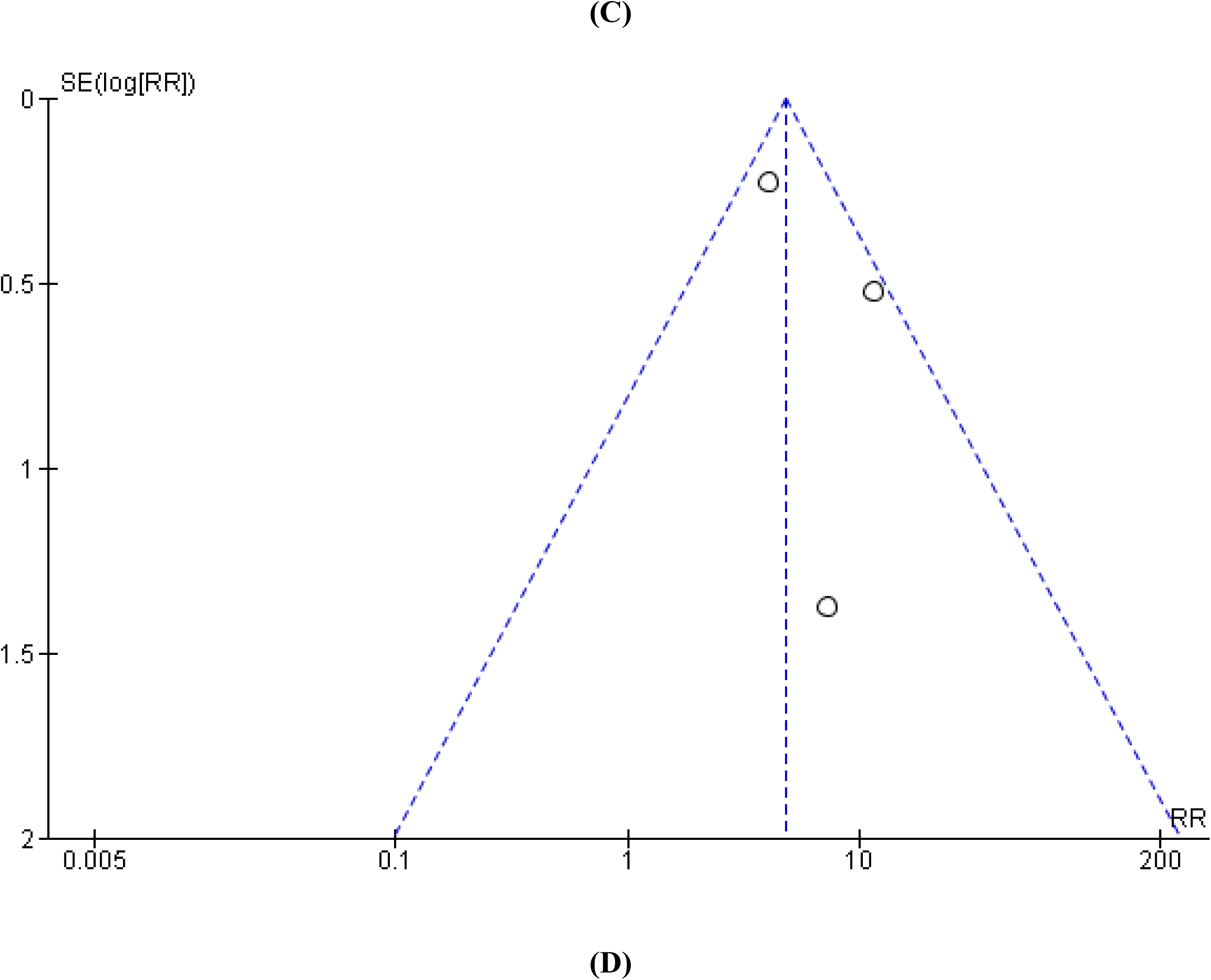
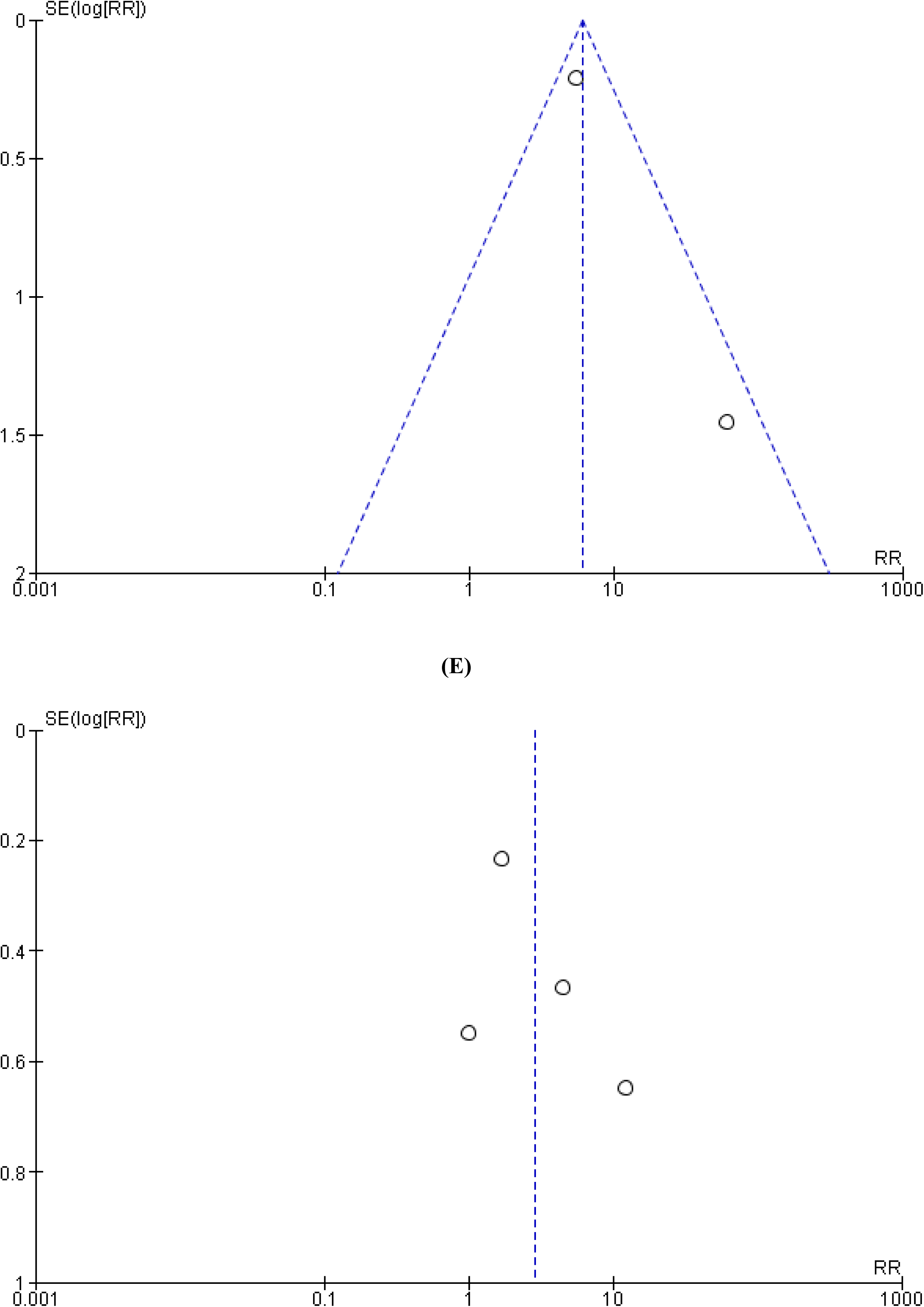
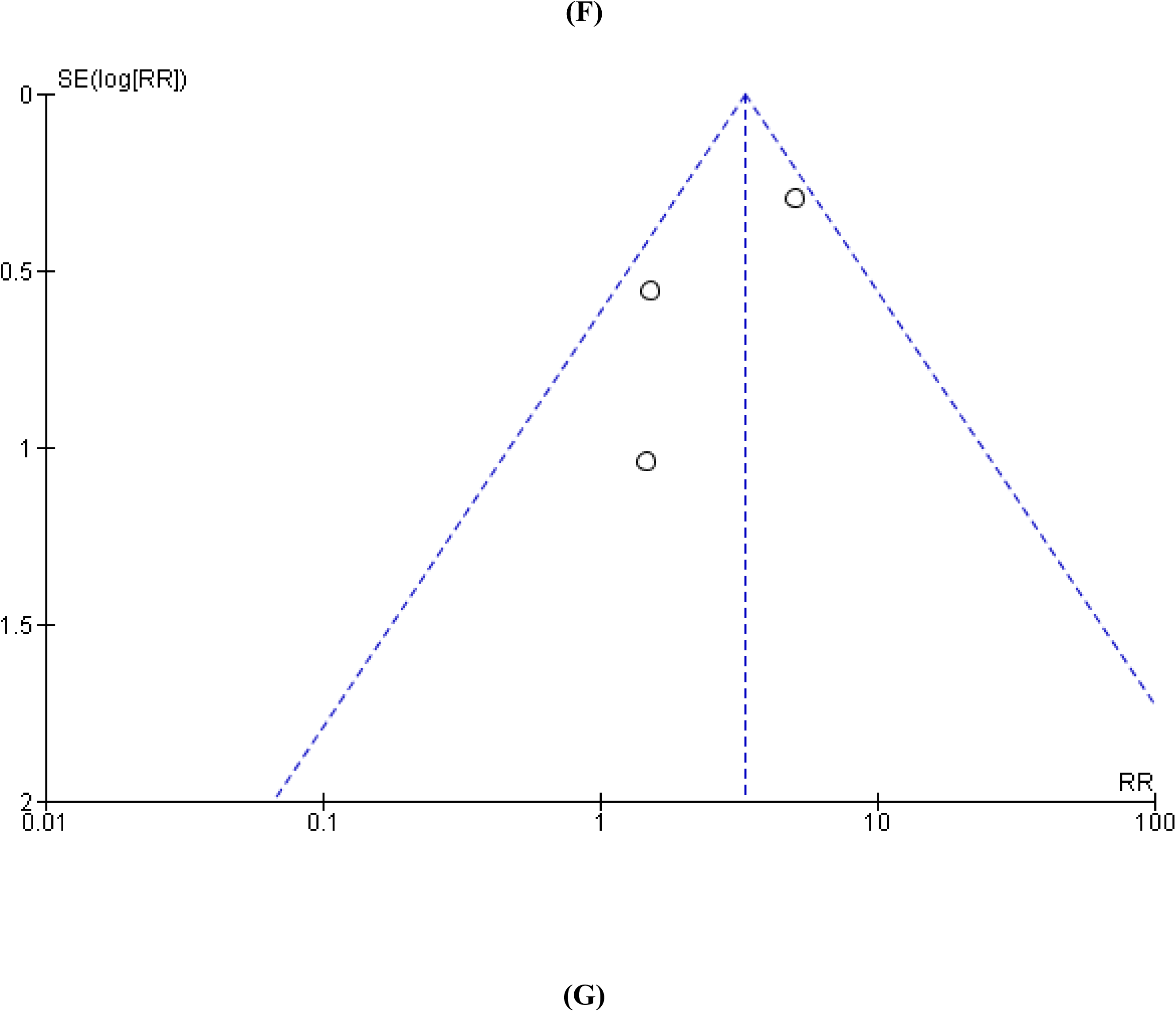
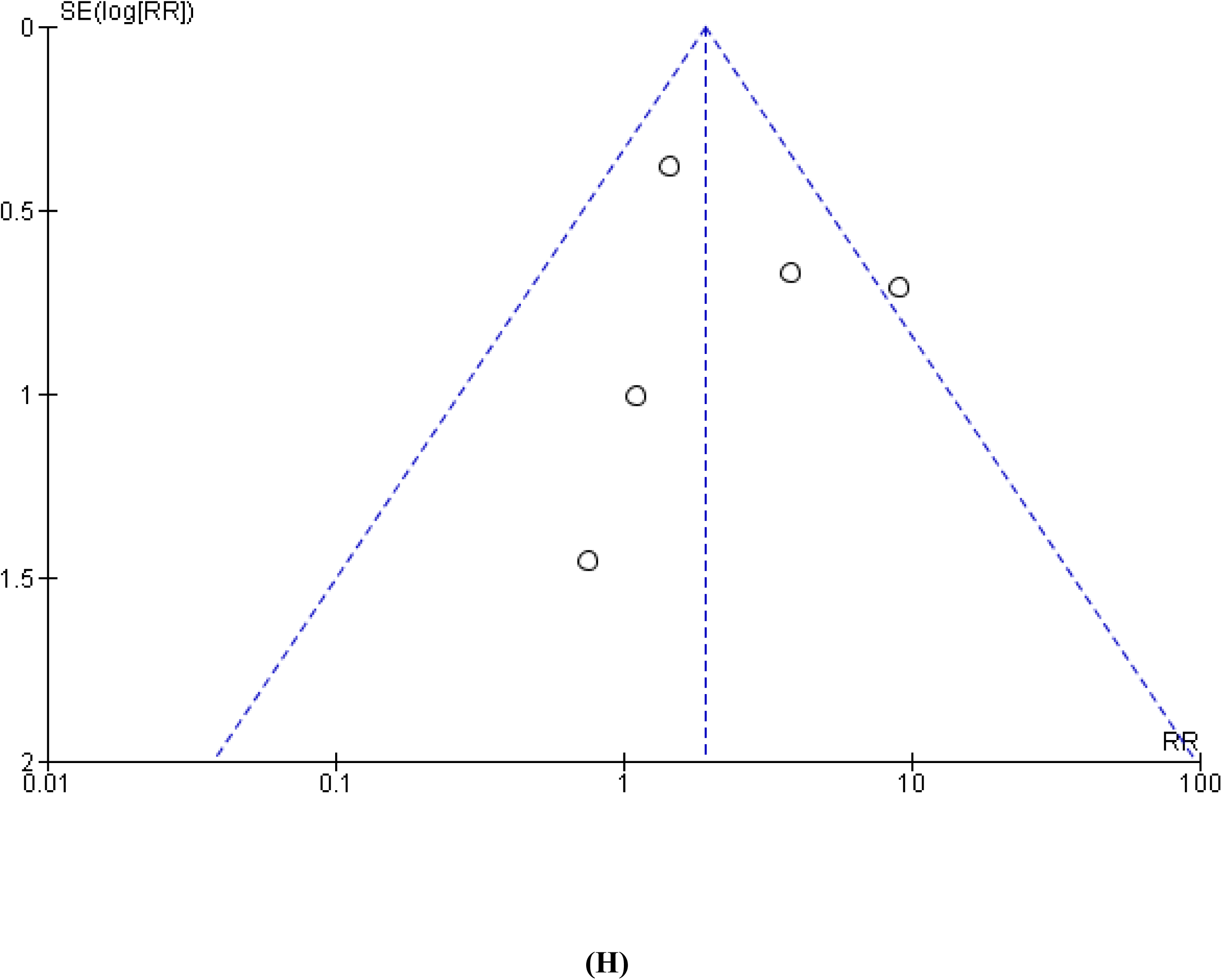
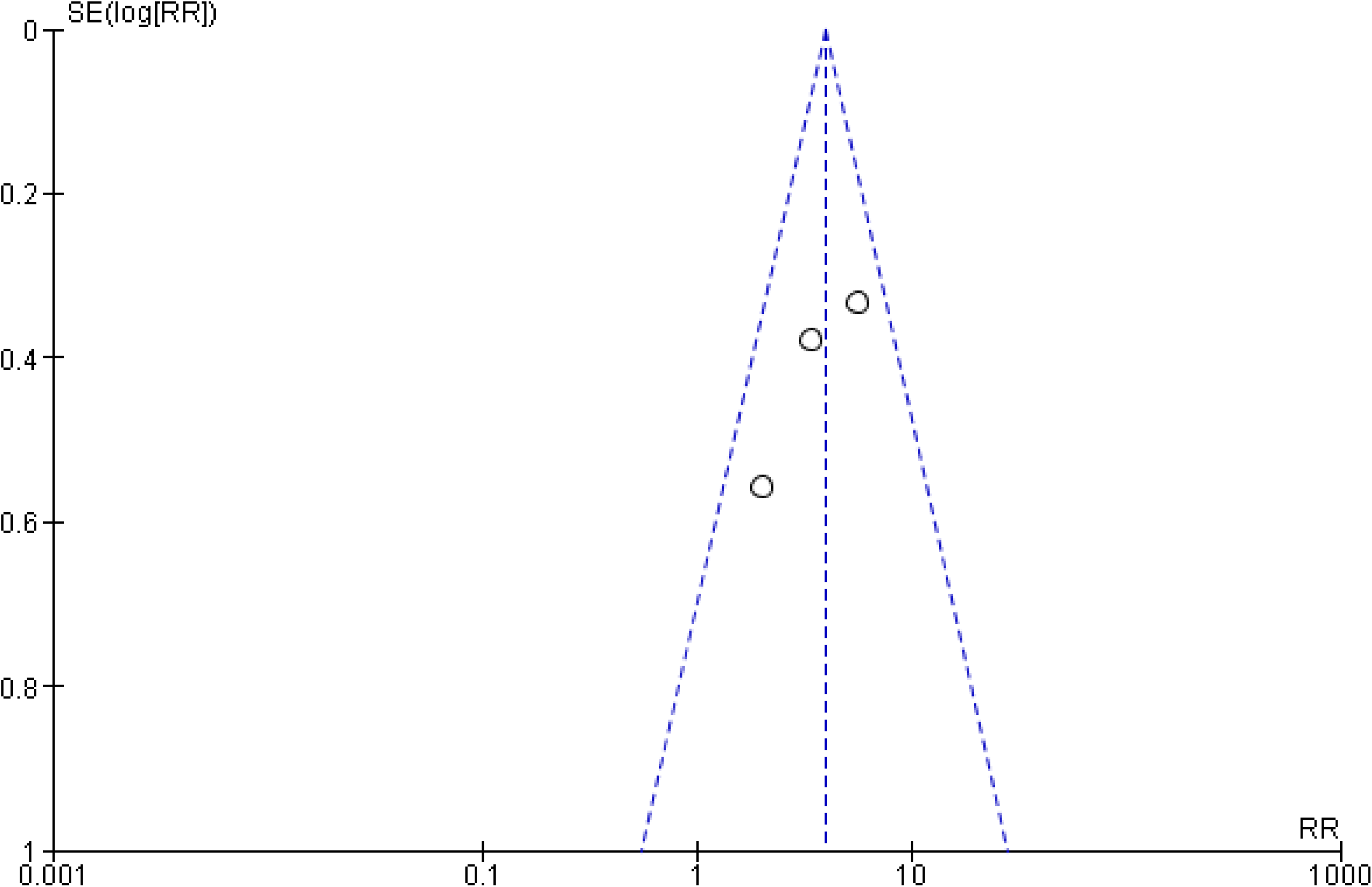
Publication bias for the outcomes of SCAD with CA vs SCAD without CA: (A) In hospital overall mortality, (B) Post-discharge overall mortality, (C) Acute heart failure, (D) Cardiogenic shock, (E) MACE (F) Recurrent MI, (G) Recurrent SCAD, (H) LVEF ≤ 40%

